# Fitbit-measured physical activity is inversely associated with incident atrial fibrillation among All of Us participants

**DOI:** 10.1101/2024.06.27.24308221

**Authors:** Souptik Barua, Dhairya Upadhyay, Aditya Surapaneni, Morgan Grams, Lior Jankelson, Sean Heffron

**Affiliations:** Division of Precision Medicine; Leon H. Charney Division of Cardiology, New York University Grossman School of Medicine, New York, NY, USA; NYU Center for the Prevention of Cardiovascular Disease, Department of Medicine, New York University Grossman School of Medicine, New York, NY, USA

## Abstract

**Background and Aims:** Individuals who report meeting weekly moderate to vigorous physical activity (MVPA) guidelines have lower risk of atrial fibrillation (AF). However existing studies have relied on subjective questionnaires or short-duration (<1 week) objective assessments using accelerometry. The objective of this research was to investigate an association between moderate to vigorous physical activity (MVPA) levels and the incidence of atrial fibrillation (AF), utilizing long-term, free-living accelerometry data.

**Methods:** 1-year Fitbit data, in addition to survey and electronic health record (EHR) data, were extracted from the NIH *All of Us* (AoU) research database. Cox proportional hazards regression was used to model the association of average MVPA (both continuous and categorized as <30, 30-150,151-300, and >300 minutes/week) and incident AF over a five-year follow-up period.

**Results:** 6086 AoU participants were included (51±16 years, 70% female, 83% White, BMI 28.5±4.9 kg/m^2^, 41±12 complete weeks of Fitbit wear). 55 individuals (0.9%) experienced incident AF in the five-year follow-up period. More time spent in MVPA was associated with lower AF risk (HR = 0.89 [0.81,0.98], p=0.02), with a step-wise reduction for 151-300 mins (p=0.07) and >300 mins (p=0.04) of weekly MVPA, respectively. In a subset of 3847 participants with genomic data, this association persisted after adjustment for AF genetic risk score.

**Conclusions:** Higher amounts of objectively measured MVPA, measured using free-living, long-term accelerometry data, were inversely associated with risk of incident AF, independent of clinical and genetic risk factors.

## INTRODUCTION

Atrial fibrillation (AF) is a highly prevalent and morbid condition. The arrhythmia is associated with substantially increased risk of stroke, heart failure and cardiovascular (CV) mortality, and afflicts approximately 1 in 200 adults – including 6% of people over the age of 65 years^1,2^. Several modifiable risk factors are consistently associated with AF, including obesity, sleep apnea, hypertension, diabetes, and physical inactivity^3^.

Physical activity (PA) has myriad health benefits – particularly related to the CV system. A wealth of clinical trial and observational data support the ability of aerobic activity of at least moderate intensity to improve CV risk factors and reduce the risk of incident CV diagnoses, including AF. To date, the data in support of these observations have largely been based on self-reported activity levels, which have limited accuracy.

Several recent studies have complemented reported activity measures with short term monitoring of participants’ exercise and activity patterns. However, the short observation periods in most studies (often ∼1 week) provide a limited snapshot of behavior patterns and may not represent usual PA amounts – as behavior may be modified due to the knowledge that the participant is being monitored. Nonetheless, among participants in the REGARDS study^4^ and UK Biobank^5^ who wore an accelerometer for up to one week, greater time spent in moderate-to-vigorous PA (MVPA) demonstrated a step-wise reduction in risk of incident AF. Notably, although not assessed in the subgroup with accelerometer data, self-reported PA was associated with reduced risk of AF only in those at intermediate or high genetic risk for the arrhythmia^2,5^. The emergence of personal accelerometer devices in recent years has provided access to extensive and reliable physical activity data in free-living individuals, yet to date, there has been minimal use of such data in studies investigating risk for developing AF.

All of Us (AoU), a prospective National Institutes of Health cohort with a particular focus on enrolling individuals from groups traditionally underrepresented in research, includes a subgroup of subjects who allowed incorporation of PA measures collected by their personal accelerometers (Fitbit). In contrast to other cohorts, these data provide extended observations of individual activity, rather than from a single, voluntary measurement period. Our goal was to investigate the influence of physical activity amount and intensity, as measured by accelerometer, on incident AF within AoU.

## METHODS

### Study Population

Data for this study came from the All of Us research program. AoU enrolled individuals aged 18 years or older at one of multiple clinical centers in the United States. The AoU Institutional Review Board provides approval for all aspects of the AoU. Complete details of the AoU research protocol have been previously published^6^. The dataset used was drawn from the AoU Controlled Tier Dataset version 7. We focused on enrolled participants who owned a Fitbit and agreed to share their Fitbit data. For these participants, in addition to Fitbit data, also available were demographics, physical measurements, vital signs and surveys collected at enrollment, and individual electronic health record (EHR) data both prior to and following AoU enrollment.

We identified 8177 AoU participants who consented to share both Fitbit and EHR data, had at least one year of Fitbit data, were 18 years or older at initiation of Fitbit wear, and did not have a prevalent atrial fibrillation diagnosis before initiation of Fitbit wear. The date range for Fitbit wear in the cohort was January 1, 2016 to June 30, 2022. Among the 8177 participants, we excluded 2091 participants with insufficient, invalid, or unusable Fitbit data, leaving a final analytic sample of 6086 participants (**Figure 1**). Details of these Fitbit-based exclusion criteria are presented in **Appendix A**.

**Figure 1:**
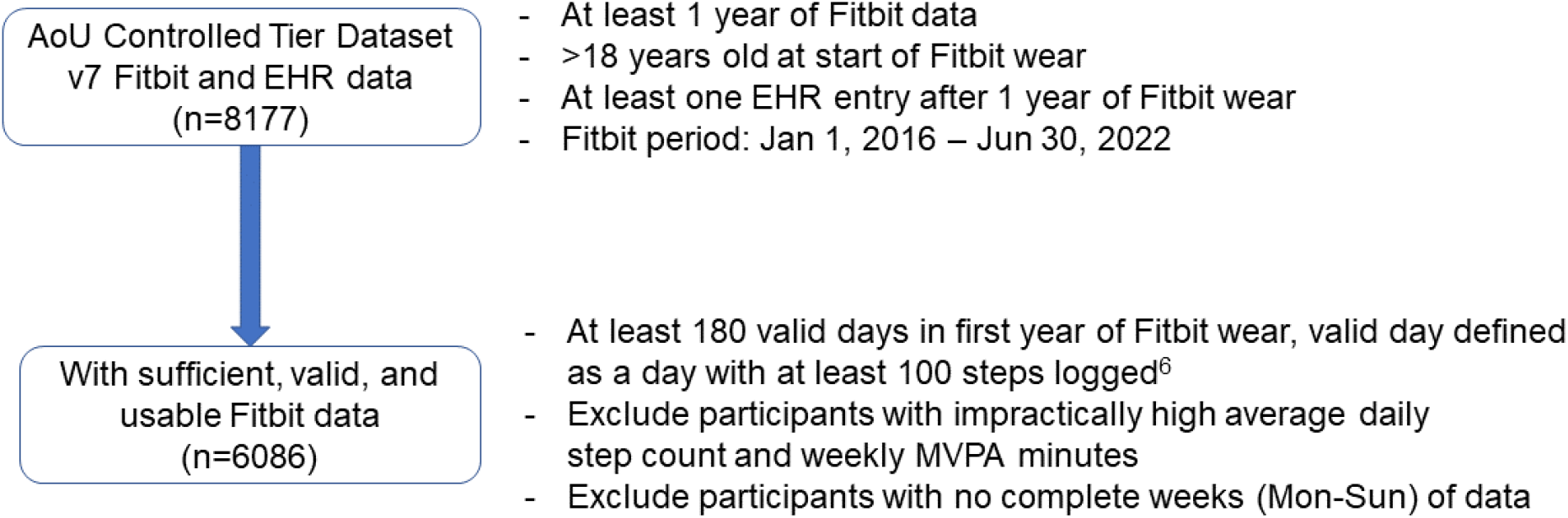
CONSORT diagram

### Fitbit data

A detailed description of AoU Fitbit data has been published previously^7^. Participants’ data had direct identifiers removed and all date/time fields were subjected to date shifting by a random number between 1 and 365 days in accordance with approved AoU privacy policies. We estimated a participant’s average weekly MVPA over the entirety of their first available full year of Fitbit data as follows: First, we computed MVPA for each day using the “fairly active” and “very active” Fitbit parameters, which have been shown to map to known definitions of moderate activity (3-6 METs in at least 10-min bouts) and vigorous activity (≥6 METs in at least 10-min bouts), respectively^8,9^. Then, we calculated MVPA mins over all valid days of Fitbit wear over the 1-year period, with a valid day defined as a day with at least 100 steps recorded^7^. Next, for complete weeks of data (weeks with 7 valid days from a Monday to the next Sunday), we calculated total weekly MVPA mins as the sum of seven daily MVPA values. Finally, we averaged a participant’s weekly MVPA mins over all complete weeks within the year to get their average weekly MVPA minutes.

### Atrial Fibrillation

The follow-up period for each participant was defined as the five-year period beginning at the completion of the first year of Fitbit wear. We used validated ICD codes (**Supplementary Table 1**) to identify incident AF diagnoses from a participant’s EHR data during their 5-year follow-up period. If a participant experienced a new AF event within the period, the time from the beginning of follow-up to date of diagnosis was recorded. If there were no AF events in the 5-year follow-up period, the participant was censored at the date of the last EHR entry, or 5 years, whichever was smaller.

### Covariates

We included a number of variables as covariates in our Cox regression models. Age at start of Fitbit wear, and self-reported sex, race, and ethnicity were extracted from demographic data captured at AoU enrollment. BMI was computed using height from EHR and the most recent weight before the start of the follow-up period. History of diabetes and hypertension were identified with a validated ICD or medication code^10^ occurring in the EHR before the start of the follow-up period (**Supplementary Table 2**). History of smoking was determined via the AoU enrollment survey question “Have you had more than 100 cigarettes in your lifetime?”

### Genetic risk score computation

A subset of 3847 out of our analytic sample of 6086 participants also had whole-genome sequence (WGS) data. From their WGS data, we computed a genetic risk score (GRS) for each individual based on a published method incorporating 24 single nucleotide polymorphisms (SNP)^11^.

### Statistical analysis

We performed Cox proportional hazards regression to model the association between average weekly MVPA and the time to first AF event. We evaluated MVPA both as a continuous and categorical variable. For categorical analysis, we stratified MVPA into four groups of average weekly MVPA: <30 minutes, 30-150 minutes, 151-300 minutes, and >300 minutes, aligned with WHO guidelines^12^. The reference group was chosen as MVPA<30 mins; the value of 30 minutes was chosen as WHO guidelines use 30-mins as the minimum unit of recommended physical activity. We adjusted our Cox models for age, self-reported sex, race, and ethnicity, BMI, history of hypertension, diabetes, and smoking. We repeated the above Cox analyses in the subset with WGS data, including GRS as an additional term in the model. We also repeated the analysis with an interaction term between GRS and MVPA category, to examine if the GRS modified the benefits of greater MVPA within our cohort. Missing data for BMI was handled using multivariate imputation by chained equations^13^. A p-value of 0.05 was set *a priori* as the criterion for statistical significance. All analyses were performed with Python software using the “scikitlearn” package^14,15^.

## RESULTS

### Participant characteristics

The study population of 6086 participants was 51±16 years old, 70% female, 83% White, and 6.3% Hispanic/Latinx (**Table 1**). Within their first year of Fitbit wear, participants on average had 41±12 complete weeks of data for estimating weekly minutes of MVPA. Amounts of weekly minutes of MVPA over the 1-year wear period exhibited a wide range among participants, with a median [IQR] weekly MVPA of 202 [105,347] minutes. 55/6086 (0.9%) participants experienced incident AF during the 5-year follow-up period. Participants with an AF diagnosis over the follow-up period were 46% female and were slightly older (63±13 years) than the sample as a whole.

**Table 1:** Cohort characteristics.

| Variable | Value |
| --- | --- |
| N: | 6086 |
| Age (years) | 50.5 ± 15.8 |
| Female sex | 4284 (70.4%) |
| Race: |  |
| --White | 5057 (83.1%) |
| --Black or African American | 319 (5.2%) |
| --Asian | 210 (3.5%) |
| --Other | 472 (7.8%) |
| Ethnicity: |  |
| --Not Hispanic or Latinx | 5605 (92.1%) |
| --Hispanic or Latinx | 384 (6.3%) |
| --Other | 97 (1.6%) |
| BMI (kg/m<sup>2</sup>) | 28.51 ± 4.92 |
| History of Hypertension: | 2086 (34.3%) |
| History of Diabetes: | 754 (12.4%) |
| Smoking: ≥100 Cigarettes Lifetime: | 1954 (32.1%) |
| Weekly minutes of MVPA |  |
| ■ Mean [SD] | 255 [208] |
| ■ Median [IQR] | 202 [105,347] |
| BMI: Body Mass Index, MVPA: Moderate to Vigorous Physical Activity, SD: Standard Deviation, IQR: Interquartile Range |  |

### Cox regression-MVPA as a continuous variable

Among demographic variables and cardiovascular risk factors, older age (in 10-year increments; HR: 1.67 [1.33,2.09], p<0.0001) and male sex (HR: 2.79 [1.58,4.94], p=0.004), but not race/ethnicity, BMI, hypertension, diabetes or tobacco history were associated with an increased likelihood of AF incidence. After adjusting for these covariates, MVPA was inversely associated with AF incidence (in one-hour increment; HR: 0.89 [0.81,0.98], p=0.02), indicating that every additional hour of MVPA was associated with an 11% reduced risk of AF (**Table 2**).

**Table 2:**
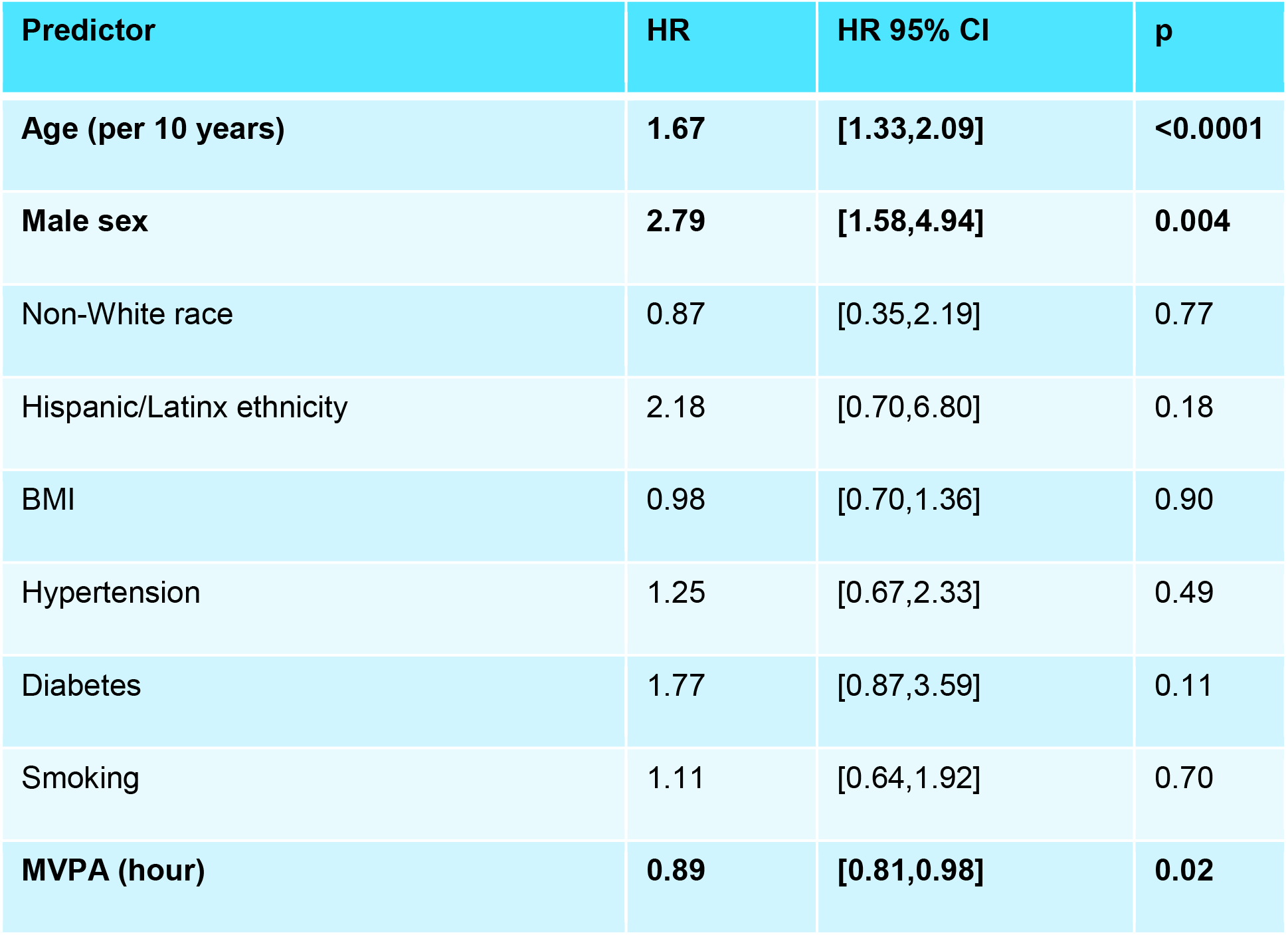
Results of Cox proportional hazards regression comparing MVPA in hours with time to an incidence of AF. MVPA: Moderate to vigorous physical activity. AF: Atrial fibrillation. (n=6086)

| Predictor | HR | HR 95% CI | p |
| --- | --- | --- | --- |
| Age (per 10 years) | 1.67 | [1.33,2.09] | <0.0001 |
| Male sex | 2.79 | [1.58,4.94] | 0.004 |
| Non-White race | 0.87 | [0.35,2.19] | 0.77 |
| Hispanic/Latinx ethnicity | 2.18 | [0.70,6.80] | 0.18 |
| BMI | 0.98 | [0.70,1.36] | 0.90 |
| Hypertension | 1.25 | [0.67,2.33] | 0.49 |
| Diabetes | 1.77 | [0.87,3.59] | 0.11 |
| Smoking | 1.11 | [0.64,1.92] | 0.70 |
| MVPA (hour) | 0.89 | [0.81,0.98] | 0.02 |

### Cox regression-MVPA as a categorical variable

We also categorized participants by average weekly MVPA based on WHO guidelines^12^: <30 minutes, 30-150 minutes, 151-300 mins, and >300 minutes. AF incidence was 1.8% (6/332) in the <30 minutes group, 0.9% (18/1949) in the 30-150 minutes group, 0.8% (14/1840) in the 151-300 minutes group, and 0.9% (17/1910) in the >300 minutes group. We observed a step-wise trend in lower risk of incident AF with increasing MVPA category (**Table 3**), with individuals who got >300 mins of MVPA per week having a significant reduction in AF risk compared to those who got <30 mins of MVPA per week (HR = 0.35 [0.13,0.95], p = 0.04) (**Figure 2**).

**Table 3:** Results of Cox proportional hazards regression comparing MVPA stratified according to WHO thresholds with time to an incidence of AF. MVPA: Moderate to vigorous physical activity. AF: Atrial fibrillation. (n=6086)

| Predictor | HR | HR 95% CI | p |
| --- | --- | --- | --- |
| Age (per 10 years) | 1.66 | [1.33,2.09] | <0.0001 |
| Male sex | 2.68 | [1.50,4.77] | 0.0008 |
| Non-White race | 0.89 | [0.35,2.19] | 0.77 |
| Hispanic/Latinx ethnicity | 2.18 | [0.70,6.80] | 0.18 |
| BMI | 0.98 | [0.70,1.36] | 0.90 |
| Hypertension | 1.25 | [0.67,2.33] | 0.49 |
| Diabetes | 1.77 | [0.87,3.59] | 0.11 |
| Smoking | 1.11 | [0.64,1.92] | 0.70 |
| **MVPA** |  |  |  |
| • 30-150 mins | 0.62 | [0.24,1.57] | 0.31 |
| • 151-300 mins | 0.40 | [0.15,1.07] | 0.07 |
| • >300 mins | 0.35 | [0.13,0.95] | 0.04 |

**Figure 2.**
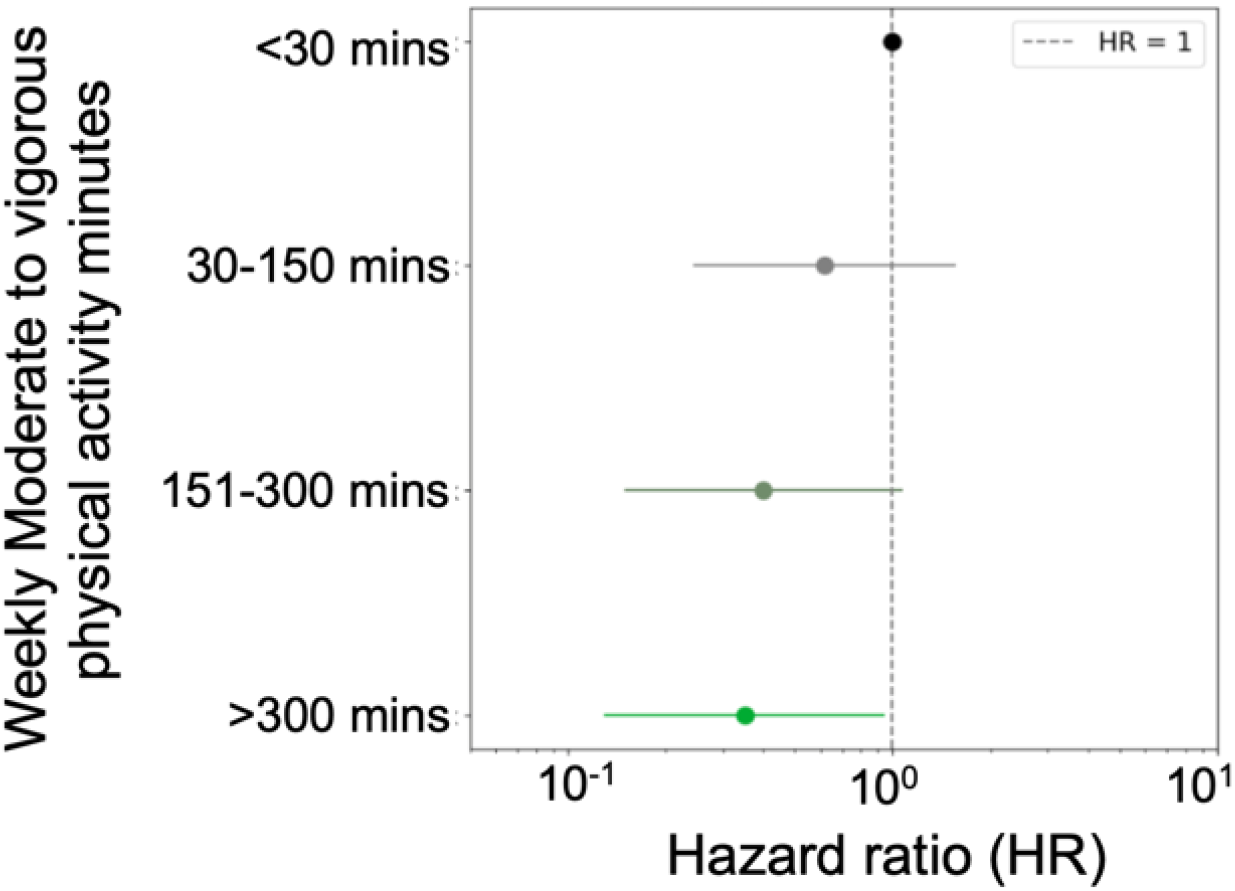
Hazard ratios for the four MVPA categories: <30 mins, 30-150 mins, 151-300 mins, and >300 mins of weekly MVPA over the 1-year Fitbit assessment.

### Genetic risk score analysis

Out of 3847 participants with available WGS data, 38 had accrued a new diagnosis of AF within the 5-year follow-up period. Characteristics of this subset of participants are detailed in **Supplemental Table 3**. The subset of individuals with and without GRS had small differences in distributions of age (51.1 vs 49.6 years old, p<0.001) and race (82.3 vs 84.5% White, p<0.05), but not sex (71.1% vs 69.1% female, p=0.11) and ethnicity (91.7% vs 92.8% Not Hispanic/Latinx, p =0.13). The number of AF events in GRS categorized by tertiles were similar in the highest (18/1258, 0.5%), intermediate (10/1275, 0.3%) and lowest (10/1276, 0.3%) GRS tertiles (χ^2^-statistic=3.49, p=0.17). Including GRS in the Cox model with continuous MVPA showed a weak association between GRS and AF incidence (HR: 1.09 [1.00,1.20], p = 0.06), with unchanged associations for other variables including MVPA (**Table 4**). This weak association held when adding GRS to the Cox model with categorical MVPA (GRS HR: 1.10 [1.00,1.21], p =0.05). Notably there was no statistical significance for any interaction term between GRS and MVPA (p=0.26, **Supplementary Table 4**), indicating higher MVPA was independently beneficial for AF risk reduction regardless of a participant’s GRS.

**Table 4:** Cox model including genetic risk score (GRS). (n=3847)

| Predictor | HR | HR 95% CI | p |
| --- | --- | --- | --- |
| **Age** | **1.54** | **[1.18,2.02]** | **0.002** |
| **Female sex** | **2.59** | **[1.31,5.13]** | **0.006** |
| Non-White race | 0.93 | [0.32,2.72] | 0.90 |
| Hispanic/Latinx ethnicity | 1.06 | [0.21,5.36] | 0.95 |
| BMI | 1.06 | [0.76,1.47] | 0.74 |
| Hypertension | 1.00 | [0.46,2.16] | 0.99 |
| Diabetes | **2.22** | **[1.02,4.86]** | **0.04** |
| Smoking | 1.11 | [0.64,1.92] | 0.70 |
| Genetic risk score (GRS) | 1.09 | [1.00,1.20] | 0.06 |
| **MVPA (hours)** | **0.87** | **[0.76,0.98]** | **0.03** |

## DISCUSSION

We report a step-wise reduction in risk of incident AF in association with accelerometry-based measurements of MVPA. To our knowledge, this is the first study to investigate MVPA estimated from free-living accelerometer data of >7 days duration with risk for AF. In a subset of participants with genomic data, this association between higher MVPA and lower AF risk persisted independently of participant’s genetic risk score.

Similar to our findings in AoU, investigators using data from participants in the UK Biobank (UKBB) who wore an accelerometer for up to one week, found that increasing time spent in moderate-to-vigorous PA demonstrated a step-wise reduction in risk of incident AF after adjustment for age, sex, race, alcohol and other CVD risk factors^5^. The step-wise association plateaued at approximately 200 minutes of MVPA weekly. In line with well-established evidence, older age and male sex were significantly associated with risk of incident AF. Notably, self-reported PA was not associated with AF risk within this sub-group with objectively measured activity, but was inversely associated with AF risk within the UKBB cohort as a whole. Further, participants in the UKBB study with accelerometer measured activity ≥150min MVPA had much lower risk for cardiovascular disease in general than those with self-reported activity ≥150min MVPA, again highlighting the greater reliability of objective MVPA assessment^16^. Self-reported PA in UKBB was associated with reduced risk of AF only in those at intermediate or high genetic risk of AF, whereas objectively measured cardiovascular fitness was strongly inversely associated with incident AF regardless of genetic risk^2,5^. Importantly, there is poor correlation between self-reported and accelerometer-measured physical activity generally^17^, including within UKBB. Another study (REGARDS) assessed accelerometer-measured PA over 4-7 days in 5147 subjects – also finding a stepwise reduction in AF risk across quartiles of moderate-to-vigorous physical activity^4^.

Our study has several limitations. MVPA estimated using a consumer device such as the Fitbit may be less accurate than research-grade actigraphy devices^18,19^. However, while studies have found differences in MVPA minutes estimated by the two devices, most studies have shown moderate to high correlation, indicating that the step-wise reduction of AF risk with increasing MVPA would hold for research-grade accelerometers albeit at different MVPA thresholds. While the AoU cohort overall is diverse (51% Non-White, 80% from groups under-represented in biomedical research^20^), 82% of the Fitbit cohort was White and 92% was non-Hispanic, likely because Fitbit data was voluntarily shared by individuals owning a personal Fitbit device. To address this issue, AoU has initiated the WEAR study to provide AoU participants with a free Fitbit. Furthermore, we did not account for differences in Fitbit models which may introduce biases in MVPA estimates^21^. Despite these limitations, the present report contributes unique, long-term, free-living PA data that support the importance of achieving recommended levels of MVPA in minimizing risk of developing AF. We further observe that this is particularly true among those at elevated genetic risk for AF. Finally, our findings highlight the important need to meet WHO MVPA guidelines (≥150 weekly minutes of MVPA) to minimize AF risk, and provides more evidence on the role of wearable devices in helping individuals stay on track to meet these guidelines.

## Data Availability

To ensure privacy of participants, data used for this study are available to approved researchers following registration, completion of ethics training and attestation of a data use agreement through the All of Us Research Workbench platform, which can be accessed via https://workbench.researchallofus.org/login.
Code used for this study can be made available to users of the All of Us Research Workbench platform by contacting our study team. Contact

## Acknowledgements

We gratefully acknowledge All of Us participants for their contributions, without whom this research would not have been possible. We also thank the National Institutes of Health’s All of Us Research Program for making available the participant data (Fitbit, EHR, and surveys) examined in this study.

## Data availability statement

To ensure privacy of participants, data used for this study are available to approved researchers following registration, completion of ethics training and attestation of a data use agreement through the All of Us Research Workbench platform, which can be accessed via https://workbench.researchallofus.org/login.

## Code availability

Code used for this study can be made available to users of the All of Us Research Workbench platform by contacting our study team. Contact

## References

1. Dai H, Zhang Q, Much AA, Maor E, Segev A, Beinart R, Adawi S, Lu Y, Bragazzi NL, Wu J. Global, regional, and national prevalence, incidence, mortality, and risk factors for atrial fibrillation, 1990-2017: results from the Global Burden of Disease Study 2017. Eur Heart J Qual Care Clin Outcomes 2021;7:574–582.

2. Elliott AD, Linz D, Mishima R, Kadhim K, Gallagher C, Middeldorp ME, Verdicchio CV, Hendriks JML, Lau DH, La Gerche A, Sanders P. Association between physical activity and risk of incident arrhythmias in 402 406 individuals: evidence from the UK Biobank cohort. Eur Heart J 2020;41:1479–1486.

3. Chung MK, Eckhardt LL, Chen LY, Ahmed HM, Gopinathannair R, Joglar JA, Noseworthy PA, Pack QR, Sanders P, Trulock KM, null null. Lifestyle and Risk Factor Modification for Reduction of Atrial Fibrillation: A Scientific Statement From the American Heart Association. Circulation 2020;141:e750–e772.

4. O’Neal WT, Bennett A, Singleton MJ, Judd SE, Howard G, Howard VJ, Hooker SP, Soliman EZ. Objectively Measured Physical Activity and the Risk of Atrial Fibrillation (from the REGARDS Study). Am J Cardiol 2020;128:107–112.

5. Khurshid S, Weng L-C, Al-Alusi MA, Halford JL, Haimovich JS, Benjamin EJ, Trinquart L, Ellinor PT, McManus DD, Lubitz SA. Accelerometer-derived physical activity and risk of atrial fibrillation. Eur Heart J 2021;42:2472–2483.

6. The “All of Us” Research Program. New England Journal of Medicine 2019;381:668–676.

7. Master H, Annis J, Huang S, Beckman JA, Ratsimbazafy F, Marginean K, Carroll R, Natarajan K, Harrell FE, Roden DM, Harris P, Brittain EL. Association of step counts over time with the risk of chronic disease in the All of Us Research Program. Nat Med 2022;28:2301–2308.

8. Ainsworth BE, Haskell WL, Leon AS, Jacobs DR, Montoye HJ, Sallis JF, Paffenbarger RS. Compendium of physical activities: classification of energy costs of human physical activities. Med Sci Sports Exerc 1993;25:71–80.

9. Leavitt MO. 2008 Physical Activity Guidelines for Americans

10. Writing Group for the CKD Prognosis Consortium. Estimated Glomerular Filtration Rate, Albuminuria, and Adverse Outcomes: An Individual-Participant Data Meta-Analysis. JAMA 2023;330:1266–1277.

11. Tikkanen E, Gustafsson S, Ingelsson E. Associations of Fitness, Physical Activity, Strength, and Genetic Risk With Cardiovascular Disease: Longitudinal Analyses in the UK Biobank Study. Circulation 2018;137:2583–2591.

12. Global Recommendations on Physical Activity for Health. Geneva: World Health Organization; 2010.

13. Buuren S van, Groothuis-Oudshoorn K. mice: Multivariate Imputation by Chained Equations in R. Journal of Statistical Software 2011;45:1–67.

14. Pedregosa F, Varoquaux G, Gramfort A, Michel V, Thirion B, Grisel O, Blondel M, Prettenhofer P, Weiss R, Dubourg V, Vanderplas J, Passos A, Cournapeau D. Scikit-learn: Machine Learning in Python. MACHINE LEARNING IN PYTHON.

15. Pedregosa F, Varoquaux G, Gramfort A, Michel V, Thirion B, Grisel O, Blondel M, Müller A, Nothman J, Louppe G, Prettenhofer P, Weiss R, Dubourg V, Vanderplas J, Passos A, Cournapeau D, Brucher M, Perrot M, Duchesnay É. Scikit-learn: Machine Learning in Python

16. Khurshid S, Weng L-C, Nauffal V, Pirruccello JP, Venn RA, Al-Alusi MA, Benjamin EJ, Ellinor PT, Lubitz SA. Wearable accelerometer-derived physical activity and incident disease. NPJ Digit Med 2022;5:131.

17. Prince SA, Adamo KB, Hamel ME, Hardt J, Connor Gorber S, Tremblay M. A comparison of direct versus self-report measures for assessing physical activity in adults: a systematic review. Int J Behav Nutr Phys Act 2008;5:56.

18. Matlary RED, Holme PA, Glosli H, Rueegg CS, Grydeland M. Comparison of free-living physical activity measurements between ActiGraph GT3X-BT and Fitbit Charge 3 in young people with haemophilia. Haemophilia 2022;28:e172–e180.

19. Semanik P, Lee J, Pellegrini CA, Song J, Dunlop DD, Chang RW. Comparison of Physical Activity Measures Derived From the Fitbit Flex and the ActiGraph GT3X+ in an Employee Population With Chronic Knee Symptoms. ACR Open Rheumatology 2020;2:48–52.

20. All of Us Research Program Investigators, Denny JC, Rutter JL, Goldstein DB, Philippakis A, Smoller JW, Jenkins G, Dishman E. The ‘All of Us’ Research Program. N Engl J Med 2019;381:668–676.

21. Diaz KM, Krupka DJ, Chang MJ, Peacock J, Ma Y, Goldsmith J, Schwartz JE, Davidson KW. Fitbit®: An accurate and reliable device for wireless physical activity tracking. International Journal of Cardiology 2015;185:138–140.

