## Supplemental data file for "Fitbit-measured physical activity is inversely associated with incident atrial fibrillation among All of Us participants"

**Appendices**

**Appendix A**

Criteria for valid Fitbit data

8177 participants were adults at start of Fitbit wear (>18 years old), had at least one EHR entry in the five-year follow-up period, and had at least one year of Fitbit data in the period Jan 1, 2016-Jun 30, 2022. The following criteria were used to identify participants whose first year of Fitbit data was sufficient, valid, and usable.

1. At least 180 valid days in first year of Fitbit wear, valid day defined as a day with at least 100 steps logged^6^ – The 100-step criterion used to define a valid day of Fitbit wear has been published previously^6^. We used the six-month (180-day) criteria to ensure sufficiently long Fitbit wear within the 1-year window, to ensure a reliable estimation of the participant’s MVPA over that year.
2. Exclude participants with no complete weeks (Mon-Sun) of data- To map MVPA estimation to real-world PA behavior, we define a complete week of data as a sequence of 7 MVPA values beginning from a Monday to the next Sunday. If any of these 7 values was from an invalid day, this week was excluded from weekly MVPA computation. Participants without a single complete week of valid days were excluded.
3. Exclude participants with impractically high average daily step count and weekly MVPA minutes- This criterion was used for infeasibly high values logged by the Fitbit due to erroneous measurement. We used standard winsorization criteria (mean±5SD) on average daily step count and weekly MVPA over 1-year Fitbit wear (excluding invalid days from step 1 above) to identify outliers.

| Criteria | Mean | SD | Lower cutoff  (mean – 5SD) | Upper cutoff  (mean + 5SD) |
| --- | --- | --- | --- | --- |
| Avg daily steps | 7973 | 4405 | 0* | 30,000 |
| Weekly MVPA | 252 | 217 | 0* | 1337 |

*Since mean <5 SD for both variables, lower cutoff is set to zero since negative values of steps and MVPA are meaningless.

**Supplementary Data**

**Supplementary Table 1**: ICD codes used to identify incident atrial fibrillation.

| ICD code type | Codes | Descriptions |
| --- | --- | --- |
| ICD10 | I48, I48.0, I48.1, I48.11, I48.19, I48.2, I48.20, I48.21, I48.3, I48.4, I48.91, I48.92 | Atrial fibrillation and flutter, Paroxysmal atrial fibrillation, Persistent atrial fibrillation, Longstanding persistent atrial fibrillation, Other persistent atrial fibrillation, Chronic atrial fibrillation, Chronic atrial fibrillation, unspecified, Permanent atrial fibrillation, Typical atrial flutter, Atypical atrial flutter, Unspecified atrial fibrillation, Unspecified atrial flutter |
| ICD9 | 427.31, 427.32 | Atrial fibrillation, Atrial flutter |

**Supplementary Table 2:** ICD and medication codes used to identify history of diabetes and hypertension

| Code type | Codes | Descriptions |
| --- | --- | --- |
| Diabetes | | |
| ICD10 | E10, E11, E13 | Type 1 diabetes mellitus, Type 2 diabetes mellitus, Other specified diabetes mellitus |
| ICD9 | 250 | Diabetes mellitus |
| ATC | A10 | Insulins and analogues, Blood glucose lowering drugs excluding insulin |
| Hypertension | | |
| ICD10 | I10, I11, I12, I13, I15 | Essential (primary) hypertension, Hypertensive heart disease, Hypertensive chronic kidney disease, Hypertensive heart and chronic kidney disease, Secondary hypertension |
| ICD9 | 401, 402, 403, 404, 405 | Essential hypertension, Hypertensive heart disease, Hypertensive chronic kidney disease, Hypertensive heart and chronic kidney disease, Secondary hypertension |
| ATC | C02, C03, C07, C08, C09 | Antihypertensives, Diuretics, Beta blocking agents, Calcium channel blockers, Agents acting on the renin-angiotensin system |

**Supplementary Table 3**: Demographic and clinical characteristics of a subset of participants with genomic data.

| **Variable** | **Value** |
| --- | --- |
| N: | 3847 |
| Age (years) | 51.1 ± 15.6 |
| Female sex | 2736 (71.1%) |
| Race: |  |
| --White | 3165 (82.3%) |
| --Black or African American | 210 (5.5%) |
| --Asian | 141 (3.7%) |
| --Other | 311 (8.1%) |
| Ethnicity: |  |
| --Not Hispanic or Latinx | 3527 (91.7%) |
| --Hispanic or Latinx | 257 (6.7%) |
| --Other | 63 (1.6%) |
| BMI (kg/m^2^) | 28.8 ± 5.6 |
| History of Hypertension: | 1664 (43.4%) |
| History of Diabetes: | 615 (16.0%) |
| Smoking: ≥100 Cigarettes Lifetime: | 1192 (31.0%) |
| Weekly minutes of MVPA   - Mean [SD] - Median [IQR] | 253 [205]  204 [106,343] |
| BMI: Body Mass Index, MVPA: Moderate to Vigorous Physical Activity, SD: Standard Deviation, IQR: Interquartile Range | |

**Supplementary Table 4**: Cox regression for participants with genomic data and including an interaction term between MVPA and GRS. MVPA: Moderate to vigorous physical activity. GRS: Genetic risk score. (n=3847)

| Predictor | HR | HR 95% CI | p |
| --- | --- | --- | --- |
| **Age (per 10 years)** | **1.55** | **[1.18, 2.04]** | **0.0015** |
| **Male sex** | **2.66** | **[1.34, 5.27]** | **0.005** |
| Non-White race | 0.83 | [0.29, 2.42] | 0.74 |
| Hispanic/Latinx ethnicity | 1.07 | [0.21, 5.42] | 0.93 |
| BMI | 1.05 | [0.76,1.46] | 0.77 |
| Hypertension | 0.99 | [0.46, 2.16] | 0.99 |
| Diabetes | 2.23 | [1.02, 4.88] | 0.04 |
| Smoking | 1.24 | [0.64, 2.39] | 0.52 |
| MVPA | 1.40 | [0.61, 3.23] | 0.42 |
| GRS | 1.17 | [1.01, 1.36] | 0.03 |
| MVPA:GRS | 0.98 | [0.95, 1.01] | 0.26 |
